# Explainable artificial intelligence for neuroimaging-based dementia diagnosis and prognosis

**DOI:** 10.1101/2025.01.13.25320382

**Authors:** Sophie A. Martin, An Zhao, Jiongqi Qu, Phoebe Imms, Andrei Irimia, Frederik Barkhof, James H. Cole, the Alzheimer’s Disease Neuroimaging Initiative

**Author notes:** Data used in preparation of this article were obtained from the Alzheimer’s Disease Neuroimaging Initiative (ADNI) database (adni.loni.usc.edu). As such, the investigators within the ADNI contributed to the design and implementation of ADNI and/or provided data but did not participate in analysis or writing of this report. A complete listing of ADNI investigators can be found at:http://adni.loni.usc.edu/wp-content/uploads/how_to_apply/ADNI_Acknowledgement_List.pdf.

## Abstract

INTRODUCTION: Artificial intelligence and neuroimaging enable accurate dementia prediction, but ‘black box’ models can be difficult to trust. Explainable artificial intelligence (XAI) describes techniques to understand model behaviour and the influence of features, however deciding which method is most appropriate is non-trivial. Vision transformers (ViT) have also gained popularity, providing a self-explainable, alternative to traditional convolutional neural networks (CNN). METHODS: We used T1-weighted MRI to train models on two tasks: Alzheimer’s disease (AD) classification (diagnosis) and predicting conversion from mild-cognitive impairment (MCI) to AD (prognosis). We compared ten XAI methods across CNN and ViT architectures. RESULTS: Models achieved balanced accuracies of 81% and 67% for diagnosis and prognosis. XAI outputs highlighted brain regions relevant to AD and contained useful information for MCI prognosis. DISCUSSION: XAI can be used to verify that models are utilising relevant features and to generate valuable measures for further analysis.

## 1 Background

Recent advancements in disease-modifying therapies highlight the growing need for effective strategies to identify individuals at risk of developing dementia. Traditional diagnostic methods rely on brain imaging, cognitive performance assessments, and long-term observations of behavioural patterns. However, the increasing availability of large research datasets has enabled the use of artificial intelligence (AI) for diagnosis and prognosis, with models demonstrating comparable and, in some cases, superior accuracy to traditional methods(1–4).

To facilitate the integration of AI tools in clinical practice, it is essential to ensure that models are safe, robust and trustworthy. Explainable AI (XAI) describes a set of techniques designed to identify which features drive a model’s predictions, allowing researchers and clinicians to verify that AI models are using relevant information and to detect potential biases. With the growing number of XAI techniques, inconsistencies in their outputs make it challenging to determine the most suitable approach(5–8). Some studies have also criticised XAI methods for their lack of robustness to noise and poor sensitivity to class-specific features(6,9,10). It is also difficult to validate their outputs in the absence of a ground-truth, which is particularly challenging in dementia research due to the complex, and heterogeneous nature of the underlying pathologies. Whilst there are ongoing efforts to improve the robustness of XAI, we sought to compare dementia-related saliency maps across several popular techniques and assess their utility in predicting future conversion to dementia.

The emergence of vision transformers (ViT) provides an attractive alternative to traditional convolutional neural networks (CNN), demonstrating superior predictive performance in many image-classification tasks(11). More importantly, ViTs are arguably inherently more interpretable(12,13). Their built-in attention mechanism explicitly captures relationships between all input regions and provides a direct way to understand their influence on the prediction. On the other hand, CNNs require post-hoc methods to generate explanations, as their use of abstract local features and hierarchical structure make it difficult to trace predictions back to specific regions in the input-space. However, to achieve their performance advantages, ViTs often require exposure to million-scale datasets which are scarce in the medical imaging domain(14). Transfer learning offers a solution to this by leveraging models that are pretrained on larger, non-medical datasets and finetuned on smaller task-specific datasets.

Several studies have explored deep learning for dementia prediction, reporting classification accuracies of approximately 70-99% for identifying Alzheimer’s disease (AD) patients from healthy controls and 60-90% for predicting mild-cognitive impairment (MCI) conversion (3,4). Although they demonstrate good performance, they often fail to assess model explanations. Furthermore, those that do often rely on a single XAI method, with comparisons of multiple methods remaining relatively underexplored in dementia applications(3). Bloch and colleagues conducted a systematic evaluation of deep learning and classical models for dementia diagnosis(15) demonstrating comparable performance between models based on volumetric features (89.6% balanced accuracy) and whole-brain neural networks (83.6% balanced accuracy). They compared the most influential brain regions identified by three XAI methods across classical and deep learning approaches. Liu and colleagues applied backpropagation to a 3D CNN trained to distinguish between healthy controls, MCI, and AD. In both cases, the authors found salient regions that are known to be relevant to dementia from existing literature, such as the left hippocampus. However, Bloch and colleagues also noted that deep learning models were more likely to rely on other regions that are less commonly associated with AD(15). With the high variability of XAI outputs across methods and the lack of consensus on the most reliable approach, it is difficult to confirm whether spurious findings are valid.

In this work, we applied transfer learning to finetune ViTs for dementia classification and compared their diagnostic and prognostic performance against a widely used CNN. We also investigated whether these fundamentally different modelling mechanisms result in distinct learned neuroanatomical patterns of dementia by comparing model explanations across the two architectures.

## 2 Methods

The study overview is depicted in Figure 1.

**Figure 1.**
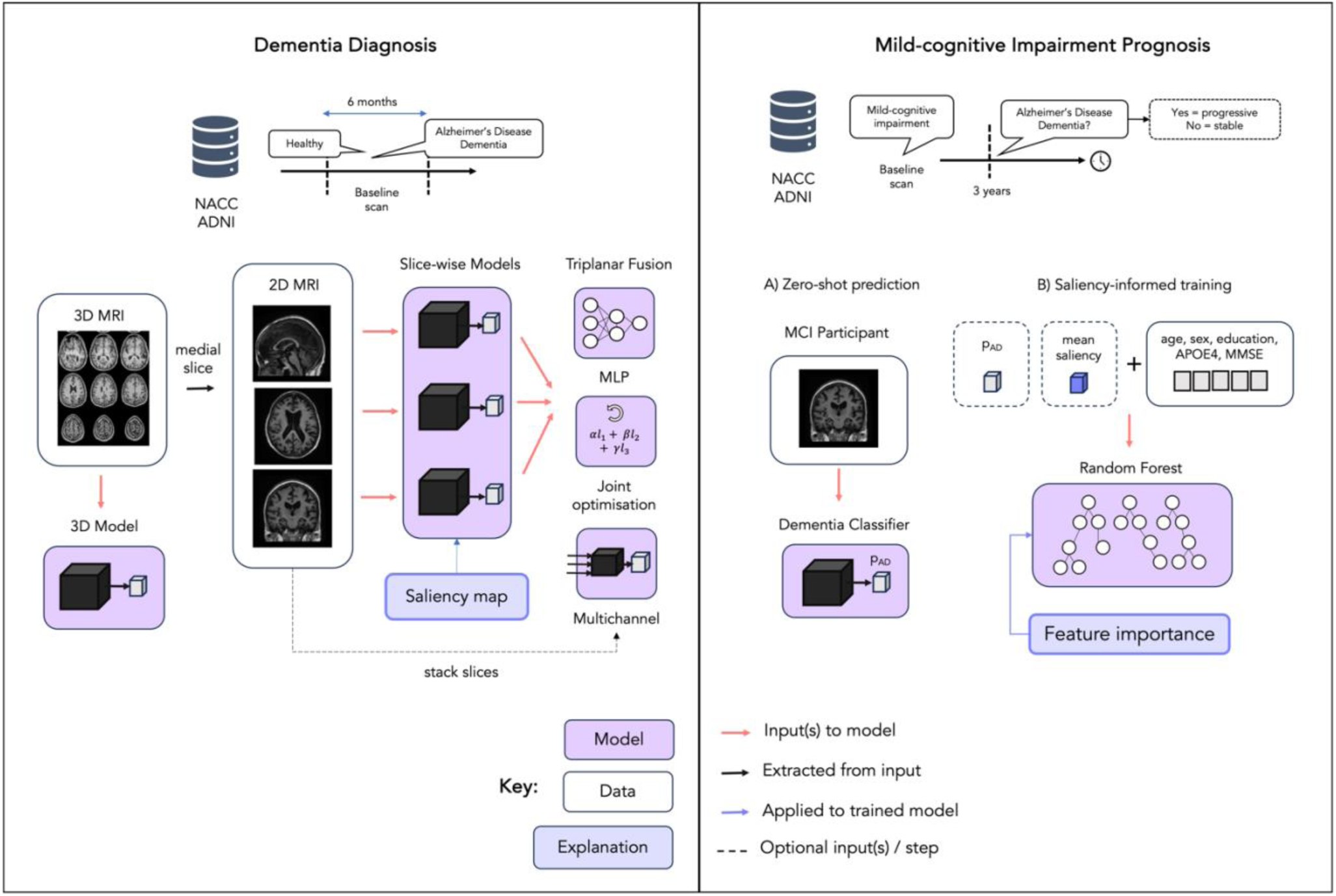
Study overview. Data from NACC was used to train and test models and ADNI was used as an external test set. Labels were assigned using baseline MRI scans and clinical diagnoses at relevant visits. For dementia classification, we applied transfer learning to finetune models on an axial, sagittal and coronal medial slice. We also trained a 3D model (ResNet) and explored the use of three triplanar fusion strategies to leverage information from each plane as an intermediary approach: i) adding a MLP layer to each output, ii) using a weighted loss to jointly optimise each network, iii) stacking slices from plane along the channel dimension. Then we applied ten explanations methods to the 2D models to generate dementia-related saliency maps. For MCI prognosis: (A) We computed the zero-shot performance of models trained on dementia classification for MCI prognosis. (B) We explored the utility of mean saliency and AD probability in a Random Forest model trained and tested on MCI prognosis. Abbreviations: MCI = mild-cognitive impairment, MMSE =mini-mental state examination, MLP = multilayer perceptron.

### 2.1 Dataset

We used demographic, clinical, and T1-weighted magnetic resonance imaging (MRI) data collated from the National Alzheimer’s Coordinating Centre (NACC) and the Alzheimer’s Disease Neuroimaging Initiative (ADNI). Models were trained on 90% of NACC participants and tested on the remaining unseen data. ADNI data was used as an external test set to assess generalisability. A summary of the demographics of each dataset is given in Table 1.

**TABLE 1:** NACC and ADNI dataset characteristics for training and test samples. Due to missing data for certain participants across the demographic features, the sample size *n*, is given for each feature and subgroup as participants with missing data have been excluded. The mean and standard deviation is given for each label respectively (AD, CN, sMCI, pMCI). Abbreviations: MMSE = mini-mental state examination, AD = Alzheimer’s disease, CN = cognitively normal, (s/p)MCI = (stable/progressive) mild-cognitive impairment. *Since ’sex’ and ’APOE4’ carriership are binary variables, we summarise these based on the proportion that are female or positive (homozygous or heterozygous) respectively. **†**Years of education.

| NACC |  |  |  |  |  |  |
| --- | --- | --- | --- | --- | --- | --- |
| Feature | Training sample |  | Test sample |  |  |  |
|  | AD/CN |  | AD/CN |  | pMCI/sMCI |  |
|  | *n* | mean±std. | *n* | mean±std. | *n* | mean±std. |
| Age | 2754 | 69.5±11.0 (73.5±9.68/68.3±11.1) | 306 | 69.0±10.3 (72.0±8.95/68.2±10.5) | 350 | 73.7±7.88 (74.7±7.84/72.6±7.80) |
| Sex* | 2754 | 0.63 (0.51/0.66) | 306 | 0.63 (0.56/0.65) | 350 | 0.43 (0.41/0.44) |
| Education** | 2750 | 15.8±2.98 (15.0±3.47/16.0±2.76) | 306 | 15.6±3.18 (14.2±3.71/15.9±2.93) | 349 | 15.6±3.16 (15.4±3.11/15.8±3.20) |
| APOE4* | 2341 | 0.40 (0.63/0.33) | 262 | 0.39 (0.55/0.35) | 343 | 0.53 (0.55/0.50) |
| MMSE | 1278 | 26.4±4.84 (20.9±5.27/28.9±1.41) | 139 | 26.3±5.20 (19.9±5.57/28.8±1.87) | 227 | 26.4 (25.7/27.3) |
| ADNI (Test sample) |  |  |  |  |  |  |
|  |  |  | AD/CN |  | pMCI/sMCI |  |
|  |  |  | *n* | mean±std. | *n* | mean±std. |
| Age |  |  | 130 | 72.8±7.45 (76.3±8.11/71.5±6.75) | 354 | 75.1±7.81 (77.4±7.29/74.3±7.83) |
| Sex* |  |  | 130 | 0.53 (0.47/0.55) | 354 | 0.40 (0.45/0.39) |
| Education† |  |  | 122 | 16.5±2.25 (15.4±2.20 / 16.9±2.13) | 354 | 16.0±2.73 (15.8±2.71/16.1±2.74) |
| APOE4* |  |  | 122 | 0.30 (0.63/0.25) | 351 | 0.31 (0.51/0.36) |
| MMSE |  |  | 107 | 27.4±2.87 (23.5±2.16/28.8±1.50) | 354 | 28.0±1.67 (27.7±1.75/28.1±1.63) |

#### 2.1.1 NACC

The NACC is a centralized data repository on Alzheimer’s disease and related dementias. It contains data from over 50,000 participants, ranging from cognitively normal individuals to those with mild cognitive impairment or dementia symptoms, with longitudinal scans and follow-up information for over 17,000 participants. Our analysis contains data from 25 centres and utilises clinical information from standardised visits conducted between September 2005 and May 2022 (June 2022 data freeze #59). Diagnoses are consensus-based, and rely primarily on clinical, cognitive and functional evaluations. Imaging data is not routinely used, as it depending on availability and site-specific protocols. The cohort used in this study consists of 3410 participants (2351 CN, 709 AD, 174 stable MCI, 176 progressive MCI). The labels are defined by the reported diagnosis at the closest visit within 6 months of the baseline scan date. For the prognostic task of predicting conversion from MCI to AD dementia, we assign stable and progressive MCI labels using baseline and follow up diagnoses. Participants who were MCI at baseline and later received an AD dementia diagnosis within a 3–4-year window were labelled as progressive MCI (pMCI). Participants who were MCI at baseline and receive an additional MCI diagnosis after 3 years or more were treated as stable MCI (sMCI).

#### 2.1.2 ADNI

The ADNI was launched in 2003 and hosts a large, dementia dataset curated from numerous research centres across the United States and Canada. The original goal was to combine imaging data, clinical and neuropsychological assessment and other biomarkers to measure the progression of MCI and early AD. Current goals include validating biomarkers for clinical trials, improving the generalizability of ADNI data by increasing diversity in the participant cohort, and providing data on the diagnosis and progression of AD. ADNI contains several imaging modalities and clinical assessments used for diagnoses such as cognitively normal, MCI and AD. ADNI incorporates rigorous image quality control procedures, including standardized protocols for MRI acquisition, preprocessing, and quality assurance to ensure consistency and reliability across sites. Participants receive a baseline visit and clinical assessment, followed by regular follow-up visits to produce a structured longitudinal database of dementia trajectories. Diagnostic labels are assigned based on expert consensus and all available participant data. In this study, we used T1-weighted MRI and diagnostic labels from a subset of 484 participants across ADNI-1, ADNI-2 and ADNI-3 cohorts (94 CN, 36 AD, 268 sMCI, 86 pMCI). Participants with a diagnosis of MCI who do not go on to receive a diagnosis of AD at any follow-up visits are labelled as sMCI. To encourage consistency with the criteria used for NACC participants, we assigned stable and progressive MCI labels according to the same criteria as NACC participants and extracted the MRI scan 3 years before the last MCI diagnosis to use for prediction. Similarly, for the pMCI group, we extracted MCI scans within a 3–4-year window before the first diagnosis of AD.

### 2.2 Data processing and quality control

For each dataset, scans were minimally pre-processed by affine registration to an asymmetric MNI-152 1x1x1mm resolution template using EasyReg(16), padded to square (or cube) and resized, followed by min-max scaling to the range 0 to 1, and normalised using the image-level mean intensity and standard deviation across the training cohort. No skull stripping was applied. For 2D models, mid-stack slices were extracted from each registered volume along three axes (axial, sagittal, coronal) and were resized to 224x224mm. For the 3D model, volumes were resized to 96x96x96mm. NACC imaging data is voluntarily provided by individual ADRCs and are not subject to a standardised imaging protocol or quality control. Therefore, these scans were visually assessed and passed through an automated image quality toolbox, MRIqc(17). Scans that failed either visual assessment or produced outliers in terms of MRIQC-derived image quality metrics (IQMs) - contrast, signal-to-noise and the coefficient of joint variation - were removed (n=158). The NACC participant inclusion criteria and data quality control procedure are detailed in Supplementary Figure A1. We also computed IQMs for ADNI scans however since ADNI has its own acquisition and quality control procedures we did not exclude scans based on these. Nonetheless, the IQM metrics highlight larger variability in image quality across the NACC dataset compared to ADNI, as shown in Supplementary Figure A2.

### 2.3 Model training

#### 2.3.1 ResNet

The ResNet model(18) is a type of CNN and a popular choice for image classification. ResNets use residual connections to give the model the flexibility to learn from previous layers and prevent issues such as vanishing gradients. ResNets consist of a series of convolutional layers, which are organized into blocks. The input image is passed sequentially through the network, allowing each convolutional layer to extract features at different resolutions via learned filters.

#### 2.3.2 Vision transformers

Vision transformers(11) are an extension of the original transformer(19) network, first developed for natural language processing, applied to image data. Unlike traditional CNNs, which process images using local filters, ViTs divide an image into patches, flatten them into one-dimensional vectors, and treat these patches as a sequence of tokens, analogous to words in text. These tokens are then processed by transformer layers, which use self-attention to capture dependencies between patches and model complex relationships within the image. The output of the final transformer layer can be linearly transformed and used for classification or other tasks.

#### 2.3.3 Is attention all you need for dementia classification?

In the natural image domain, ViTs have demonstrated superior performance over CNNs, particularly when pre-trained on large amounts of data(11). A study by Dosovitsky and colleagues suggested that pretraining datasets of approximately 100 million samples are needed to observe performance benefits of ViTs over CNNs, due in part to the need to learn spatial inductive biases(11,14). Whilst there have been emerging strategies to address this such as data augmentation and strong regularisation(20–22), many studies have found that in small-data settings such as medical imaging, ViTs can be difficult to effectively and efficiently train. Nonetheless, there have been several efforts to pretrain ViTs from scratch using 3D MRI data, before finetuning them on dementia classification tasks. For example, Dhinagar and colleagues(23) propose a scaled-down Neuroimage Transformer (NiT) that takes 3D voxels as inputs instead of patches. Under this framework, the NiT is pretrained on sex classification using data comprising of approximately 140,000 3D volumes across two datasets, UK Biobank (UKB)(24) and the 100,000 Latent Diffusion Model (LDM) synthetic dataset. On dementia classification this model was able to achieve an area-under-the-receiving-operator-curve value of 89.2%, when tested on ADNI and classifying AD patients from healthy controls. This was shown to produce an 8.9% improvement over the ViT-B-16 model with 88M parameters. Alternatively, a recent study by Kunanbayev and colleagues empirically compared several training strategies and a range of imaging datasets for pretraining, to optimise AD classification(25), reporting an optimal cross-validation accuracies of 79.6% and 81.2% in ADNI-1 and ADNI-2 respectively. However, in both cases the ViT performance was not benchmarked against a CNN for a direct comparison.

On the other hand, studies have also shown that performance gains over CNNs can be obtained through hybrid architectures(26–29). Typically, this involves adding a feature extractor network layer before feeding patches to the transformer encoder, instead of raw patches from the original input. This allows for useful features to be extracted first, allowing the ViT to converge and optimise more efficiently on smaller datasets. In this regime, Li and colleagues report balanced accuracies of 88.4% and 83.7% for the Trans-ResNet (hybrid) and ResNet respectively when finetuned on ADNI and tested on AIBL, an external dataset for AD versus control classification. Notably, these models were pretrained on brain age estimation using approximately 36,000 3D MRI volumes from UKB, highlighting the potential to effectively train ViT-based models on medical imaging data. However, in this study we have chosen not consider hybrid models to maintain the distinction between a convolution-free (ViT) and convolution-based (ResNet) model and retain the advantage of ViTs’ inherent explainability.

#### 2.3.4 Transfer learning

At the time of writing, well-established pretrained 3D ViT models have not been made publicly available for finetuning on smaller datasets. Therefore, we leveraged pretrained weights which outperform CNNs in the natural image domain and devise a framework to finetune models on 2D slices from 3D MRI volumes. This allowed us to compare the performance of a ResNet and ViT model for dementia prediction tasks and compare the explanations produced by post-hoc methods across both architectures. Models were trained on data from the NACC and ADNI. For both datasets, 90% was used for development and 10% was set aside as a hold-out test set. To leverage pretrained 2D model weights(30), we applied transfer learning and finetuned each model using mid-stack axial, sagittal and coronal slices. Both models were initially trained on ImageNet-1k(31) allowing the models to learn high-level, generalisable patterns. To finetune models effectively, we began by performing a hyperparameter search to identify the optimal training configuration for each architecture (using axial slices) and the 3D ResNet. The optimal configuration was then kept fixed throughout. We allowed all weights to be updated during finetuning. Details on the model architectures, hyperparameter search space, optimal parameters and model training details are given in Supplementary Table A8.

The optimal hyperparameters were used to obtain three plane-specific models across the two architectures: -axial, -sagittal, -coronal. Early stopping was used for each fold to reduce overfitting (patience of 10 based on the validation area-under-the-receiver-operator curve (AUROC)). We also trained a 3D ResNet-18 from scratch to benchmark against the 2D models.

#### 2.3.5 Triplanar models

Many existing studies have shown that 3D models outperform their 2D counterparts due to the ability to leverage contextual information across the volume(3,32). This is particularly relevant for neuroimaging data, as biologically relevant information is often present throughout the brain volume, rather than a single slice(32). Due to the limited availability of pretrained 3D ViT models, here we limited our focus to models trained on 2D slices.

However, as an alternative we explored fusion strategies to combine information across three orthogonal views: axial, sagittal and coronal. First, we trained an ensemble model which combines the predictions of three independent single-plane models in a fully connected feed-forward MLP layer, typically referred to as late fusion. We also explored a stacking strategy, where the three views were stacked along the channel dimension, allowing information from each slice to be utilised by a 2D model. Thirdly, we implemented a joint optimisation strategy in which the independent model weights are learned via a weighted loss, *L*, given by:

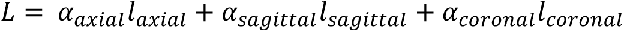

where α are factors optimised during training using a binary cross-entropy loss and training procedure described above. We also computed the average (T-AVG) which served a baseline ensemble approach.

### 2.4 Explainable artificial intelligence methods

We primarily used the captum(33) library to implement ten popular explanation methods for the main model architectures. This included model-agnostic methods such as occlusion, SHAP and LIME. We also use this library to apply neural-network-based methods such as Backpropagation, Integrated Gradients, InputXGradient, (guided) Grad-CAM and LRP. These methods all rely on access to the gradients learned during training but vary in their mechanisms for propagating values into the input image space. However, some of these methods (e.g., GradCAM) cannot be applied to ViTs or ResNets naively because they were originally designed for CNNs. Therefore, we use an alternative implementation which is model-agnostic to generate maps for ViTs and ResNets(34). Similarly, to apply the LRP method ResNets, care must be taken to account for skip connections which is not in the original design or captum implementation. Therefore, we adapted code from Otsuki and colleagues(35) to apply LRP to the ResNet architecture. For ViT models, we also used attention rollout to generate model-specific explanations. Attention rollout iteratively multiplies the attention weights from all layers, effectively tracing how information is propagated from the input image through the model. This can be extended further by imposing rules to mimic the relevance condition of LRP(36) but is not implemented in this work. Additionally, Byun and colleagues recently proposed ViT-ReciproCAM(37) as a ViT specific alternative to ReciproCAM(38) and showed that it outperforms other methods based on the ADCC metric defined by Poppi and colleagues(39). This method also has the advantage of not requiring access to the model gradients like model-agnostic approaches, therefore supporting its utility in inference-only settings, which could be required due to privacy concerns in medical contexts. Heatmaps were thresholded based on their intensities to identify the most salient regions (top 10% percentile) and reduce noise. We also applied a RELU operation to focus on positive values and scaled each map by the maximum value such that the intensities range from 0 to 1.

#### 2.4.1 Comparing model explanations

Our work seeks to understand whether brain-relevant information is being used to inform predictions as a sanity check and to identify learned biases. We devised a check to assess the amount of relevance assigned to brain and non-brain regions across the different XAI methods as a crude measure of brain-relevant specificity.

To assess whether model explanations are specific to AD, we generated group-level saliency maps by averaging the outputs across methods for different subgroups (true positives + false positives = ‘AD’, true negatives + false negatives = ‘CN’). These tests aim to shed light on the variability and specificity of XAI outputs across different techniques. However, it is important to note that a true evaluation of their utility is difficult due to the lack of individual level ground-truth references. Whilst group-level saliency maps can be compared to known biologically relevant ROIs, individual-level validation is more complex due to the heterogeneous atrophy patterns across the AD continuum, some of which can be captured by normative models(40). Additionally, we assessed the agreement of the most salient features across the different methods and architectures to identify the methods that are most robust. We computed the dice coefficient based on the most salient regions (top 10% intensity percentile) of the average ‘AD’ heatmap across the different models and methods. The dice coefficient is given by,

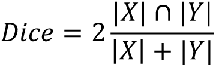

where |*X*| and |*Y*| correspond to binary maps and ∩ refers to their overlap.

#### 2.4.2 Explainability as a feature

To assess the utility of the model explanations as a predictive feature we performed a sub-analysis using mean saliency in a Random Forest model trained specifically on prognosis (i.e., classifying pMCI from sMCI). First, we used the trained dementia classifier to obtain a ‘dementia’ heatmap for each MCI participant. To reduce noise, we removed non-brain pixels using an MNI-152 brain segmentation mask and applied RELU and normalisation as before. The average intensity was included as a feature in a Random Forest classifier alongside relevant clinical features (age, sex, education, APOE4 status, MMSE score) and optionally, the zero-shot predicted probability, p_AD_ based on the MRI. An optimal configuration was trained using the scikit-learn 5-fold grid search procedure based on the average precision (AUPRC) value based on a single model and kept fixed for the rest of the analysis. Hyperparameters and the range of tested values are given in Supplementary Table A7. For this analysis, we used 75% of MCI participants for training and tested on the remaining 25%. After removing participants with missing features, this gave a training sample of 164 and test sample of 56 participants. We benchmarked the addition of saliency against two baseline models: one that used clinical and demographic features only and another with p_AD_ as feature. To capture the variability in the trained models, we report the average and standard deviation across 25 repeats for each input configuration (various random seeds).

Furthermore, once the optimal combination of saliency features and probabilities was found we applied SHAP using the captum library to identify the influence of each feature.

## 3 Results

### 3.1. Classification performance

Figures 2a and 2b demonstrate the receiver-operator curve (ROC) performance for ResNet and ViT models trained and tested on the NACC dataset, with AUC (area under curve) values shown in the legend for the diagnosis and prognosis tasks respectively. These results are summarised in Table 2.

**Figure 2.**
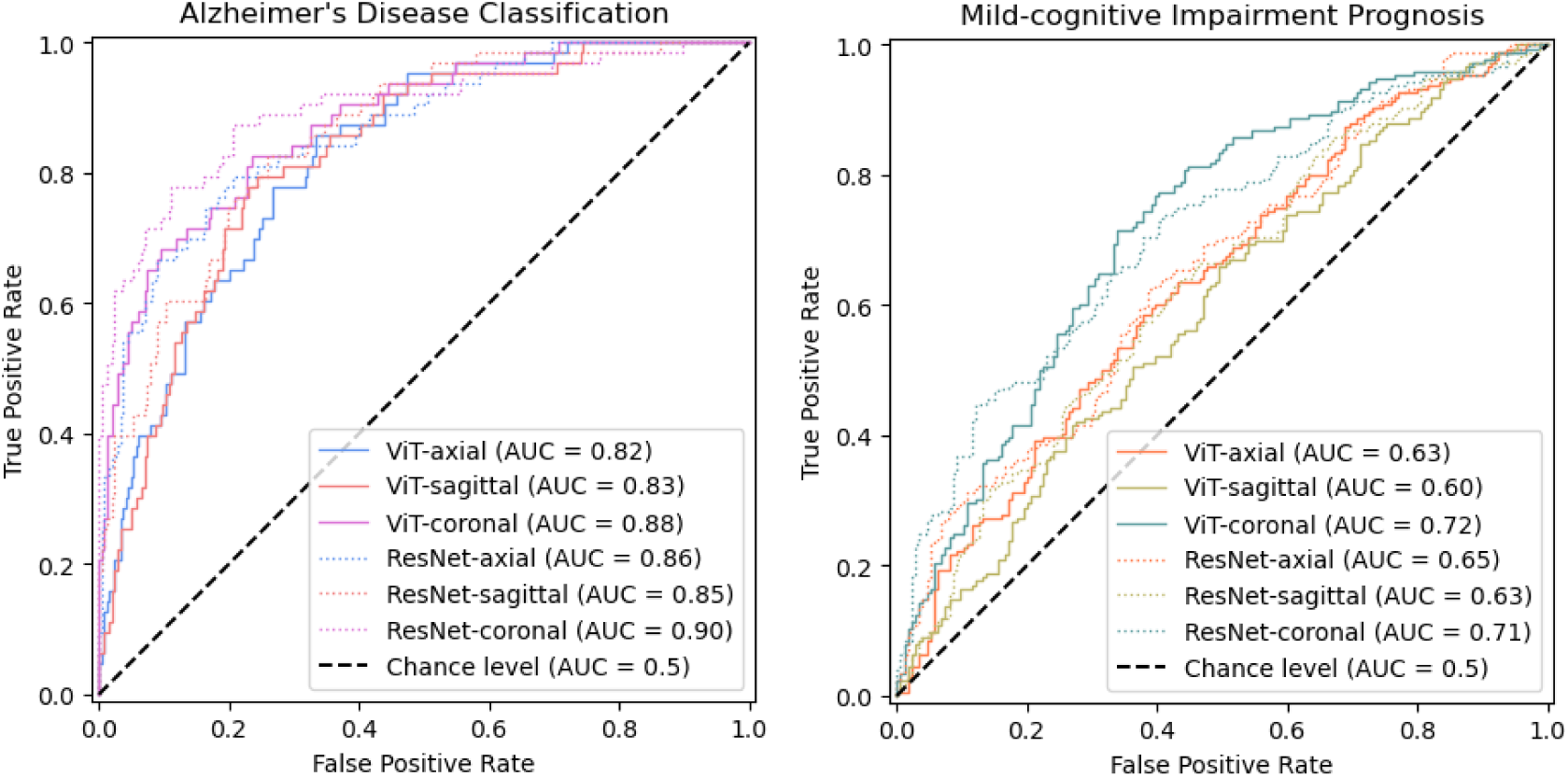
A) Receiver-operator curves for single-slice ResNet and vision transformer models trained and tested on NACC data for dementia classification (AD vs HC). Colours correspond to the different views - sky-blue: axial, pink: coronal, red: sagittal. B) Receiver-operator curves for single-slice ResNet and vision transformer models tested (zero-shot) on MCI prognosis (sMCI vs pMCI). Colours correspond to the different views-blue-green: axial, orange: coronal, yellow: sagittal.

**Figure 3.**
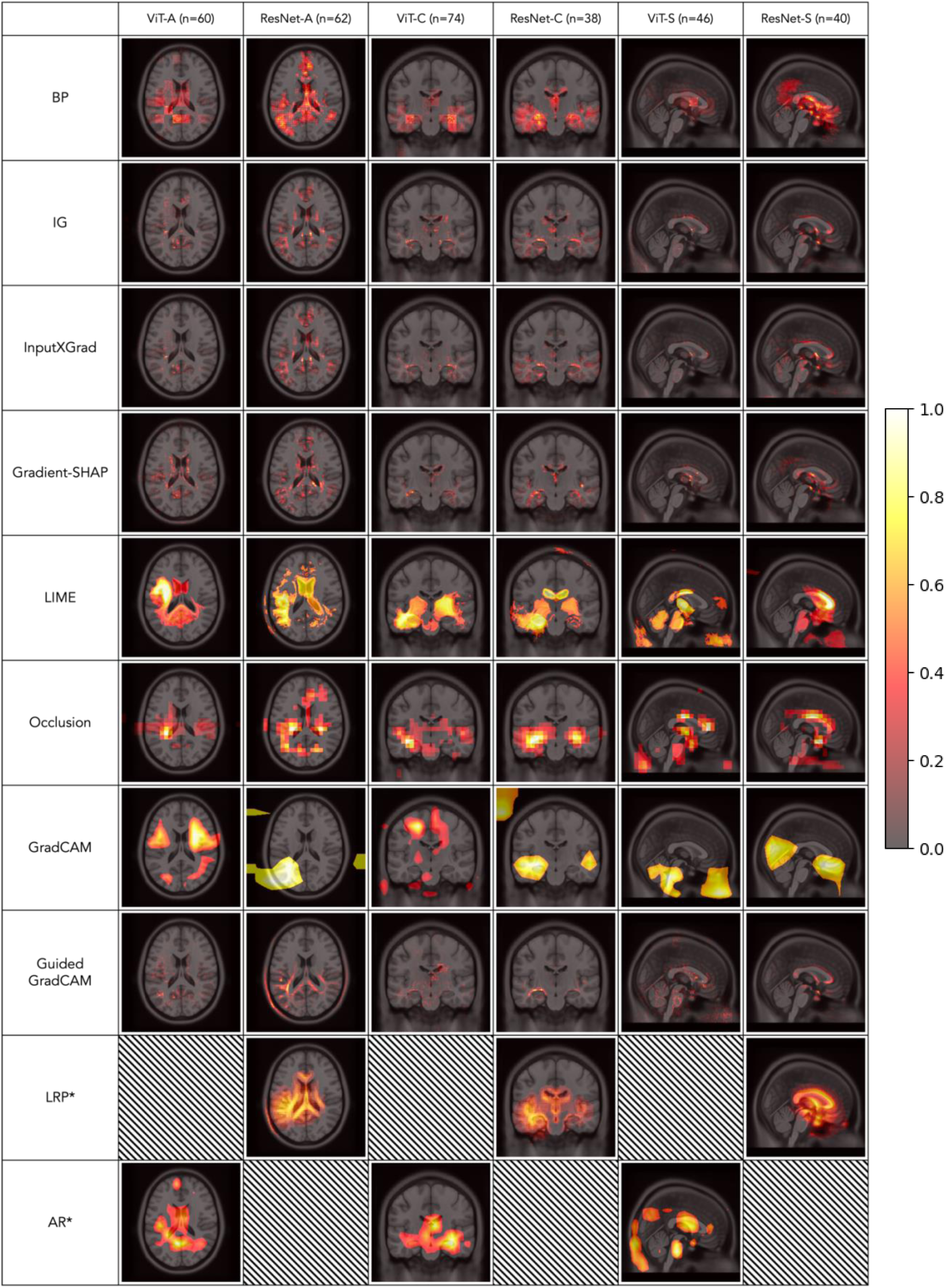
op 10% of salient pixels/regions for saliency maps generated by ten XAI methods (averaged over positive AD model predictions (TP + FP) with the corresponding number of participants given in brackets). Abbreviations: ViT = vision transformer, -A = axial, -S = sagittal, -C = coronal, LRP = layer wise relevance propagation, AR = attention rollout, BP = backpropagation, IG = integrated gradients, INPUTGRAD = InputxGradient, CAM = class activation mapping, LIME = local interpretable model explanations, SHAP = (Shap)ley values. * Denotes methods that were not implementable for both architectures.

**Figure 4.**
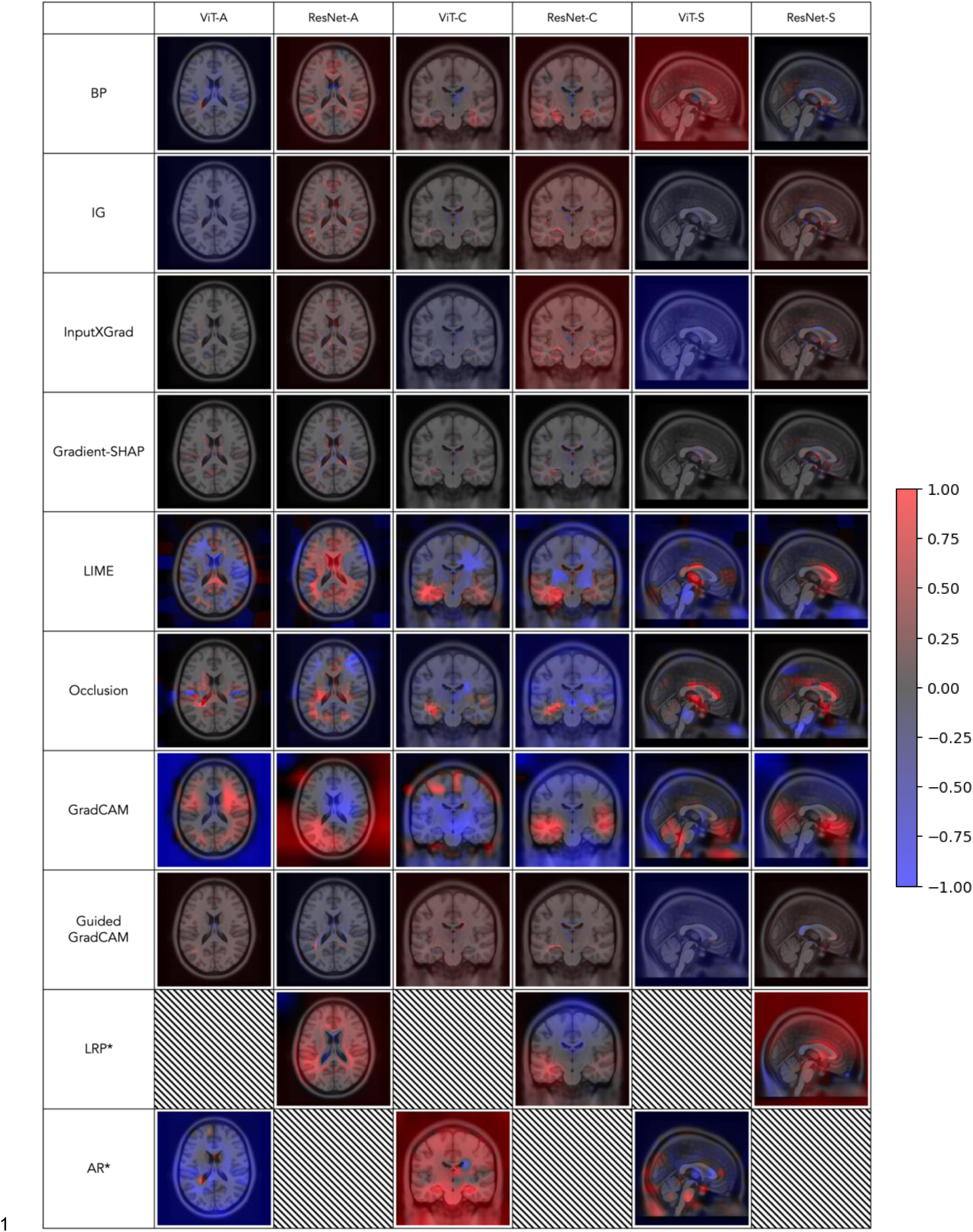
Difference between mean ‘AD’ (true positive and false positive predictions) and ‘CN’ (true negative and false negative predictions) saliency maps generated by ten XAI methods. No thresholding was applied. Abbreviations: ViT = vision transformer, -A = axial, -S = sagittal, -C.= coronal, LRP = layer wise relevance propagation, AR = attention rollout, BP = backpropagation, IG = integrated gradients, INPUTGRAD = InputxGradient, CAM = class activation mapping, LIME = local interpretable model explanations, SHAP = (Shap)ley values. * Denotes methods that were not implementable for both architectures.

#### 3.1.1 Triplanar fusion

To leverage information from orthogonal views of the medial slice, we compared three strategies: stacked, joint and late fusion. Of the three methods, joint fusion led to the best AUPRC, most evidently when using ResNet backbones for prognosis. For this task, the joint model even exceeded the performance of the fully-3D architecture.

#### 3.1.2 Cross-cohort generalisability

To assess the generalisability of these results we also evaluated the cross-cohort performance by evaluating the models on data from the ADNI. This is summarised in Table 4.

**Table 3.** Performance of single slice and triplanar models for two tasks, diagnosis and prognosis (zero-shot) across both architectures in a held-out test set from the NACC data. Abbreviations: BACC = balanced accuracy, AUROC = area-under-the-receiver-operator-curve, AUPRC = area-under-the-precision-recall-curve, AD = Alzheimer’s disease, CN = cognitively normal, (s/p)MCI = (stable/progressive) mild-cognitive impairment.

| NACC Test Performance |  |  |  |  |  |  |  |  |  |  |  |  |  |
| --- | --- | --- | --- | --- | --- | --- | --- | --- | --- | --- | --- | --- | --- |
|  |  | Diagnosis (AD (63) vs. CN (243)) |  |  |  |  |  | Prognosis (sMCI (174) vs. pMCI (176)) |  |  |  |  |  |
|  |  | Vision Transformer |  |  | ResNet |  |  | Vision Transformer |  |  | ResNet |  |  |
|  |  | BACC | AUROC | AUPRC | BACC | AUROC | AUPRC | BACC | AUROC | AUPRC | BACC | AUROC | AUPRC |
| Single slice | Axial | 68.6 | 82.5 | 55.0 | 77.2 | 86.5 | 70.0 | 56.7 | 63.4 | 61.2 | 56.3 | 65.1 | 66.0 |
|  | Coronal | 78.7 | 87.8 | 72.5 | 75.2 | 89.7 | 80.4 | 63.8 | 72.1 | 69.9 | 62.1 | 71.4 | 72.6 |
|  | Sagittal | 65.9 | 82.6 | 54.3 | 68.8 | 85.4 | 65.9 | 56.1 | 59.6 | 58.5 | 56.2 | 63.2 | 61.5 |
| Triplanar | Stacked | 77.8 | 88.5 | 72.2 | 81.1 | 90.5 | 79.7 | 60.6 | 64.6 | 62.8 | 65.0 | 72.0 | 71.6 |
|  | Joint | 78.6 | 89.1 | 73.2 | 80.7 | **91.7** | **82.2** | 63.6 | 70.4 | 69.9 | 67.0 | **73.4** | **73.1** |
|  | Late | 74.7 | 87.6 | 67.9 | 78.5 | 90.5 | 80.2 | 61.8 | 70.3 | 67.3 | **67.3** | 72.6 | 72.2 |
|  | 3D | - | - | - | **83.4** | 89.5 | 82.1 | - | - | - | 65.7 | 72.7 | 73.0 |

**Table 4.** Zero-shot, cross-cohort generalisability of NACC-trained single slice and triplanar models for two tasks, diagnosis and prognosis across both architectures in a held-out test set from the ADNI data. Abbreviations: BACC = balanced accuracy, AUROC = area-under-the-receiver-operator-curve, AUPRC = area-under-the-precision-recall-curve, AD = Alzheimer’s disease, CN = cognitively normal, (s/p)MCI = (stable/progressive) mild-cognitive impairment.

| ADNI Test Performance |  |  |  |  |  |  |  |  |  |  |  |  |  |
| --- | --- | --- | --- | --- | --- | --- | --- | --- | --- | --- | --- | --- | --- |
|  |  | Diagnosis (AD (36) vs. CN (94)) |  |  |  |  |  | Prognosis (sMCI (286) vs. pMCI (86)) |  |  |  |  |  |
|  |  | Vision Transformer |  |  | ResNet |  |  | Vision Transformer |  |  | ResNet |  |  |
|  |  | BACC | AUROC | AUPRC | BACC | AUROC | AUPRC | BACC | AUROC | AUPRC | BACC | AUROC | AUPRC |
| Single slice | Axial | 75.8 | 84.4 | 66.8 | 70.2 | 84.6 | 69.1 | 60.9 | 64.0 | 36.6 | 65.7 | 68.1 | 40.9 |
|  | Coronal | 80.3 | 90.0 | 85.1 | 74.3 | 93.2 | 86.3 | 65.4 | 71.7 | 44.8 | 58.4 | 72.6 | 44.0 |
|  | Sagittal | 65.4 | 77.6 | 55.9 | 76.0 | 87.4 | 75.9 | 56.5 | 62.9 | 33.1 | 57.9 | 65.8 | 34.4 |
| Triplanar | Stacked | 78.1 | 87.9 | 80.6 | 81.7 | 91.3 | 85.9 | 65.1 | 69.4 | 41.1 | 62.8 | 72.6 | 45.6 |
|  | Joint | 77.2 | 88.8 | 79.4 | 81.5 | 93.0 | 86.2 | 63.6 | 69.1 | 39.2 | 67.9 | **75.6** | **48.5** |
|  | Late | 78.9 | 89.1 | 82.2 | 79.6 | 91.1 | 84.1 | 64.5 | 70.6 | 43.1 | 64.8 | 73.3 | 48.1 |
|  | 3D | - | - | - | **85.4** | **94.6** | **89.6** | - | - | - | **69.4** | 73.4 | 46.1 |

### 3.2 Saliency maps for dementia diagnosis

We applied ten distinct explanation techniques to explore which regions the model is using to inform its prediction. First, we explored this on a group-level by producing average ‘AD’ heatmaps based on the model’s true and false positive predictions, thresholded to identify the most salient pixels (Figure 3). As a sanity check we assessed whether these lie within the brain, both visually and by computing the proportion of non-zero pixels (Supplementary Figure A4). Across the methods, we observe high saliency in regions such as the ventricles, hippocampus, corpus callosum and cingulate gyrus. To quantify consistency between model architectures, we computed the pairwise dice overlap of the most salient regions for each plane (axial, sagittal or coronal slices – see Figure 5). XAI methods differ in their ability to distinguish between positively and negatively-affecting contributions, however, to capture their group-level sensitivity, we compute the difference in the average ‘AD’ and average ‘CN’ maps in Figure 6 (no thresholding applied). Average ‘CN’ heatmaps based on true and false negative predictions are shown in Supplementary Figure A5.

**Figure 5.**
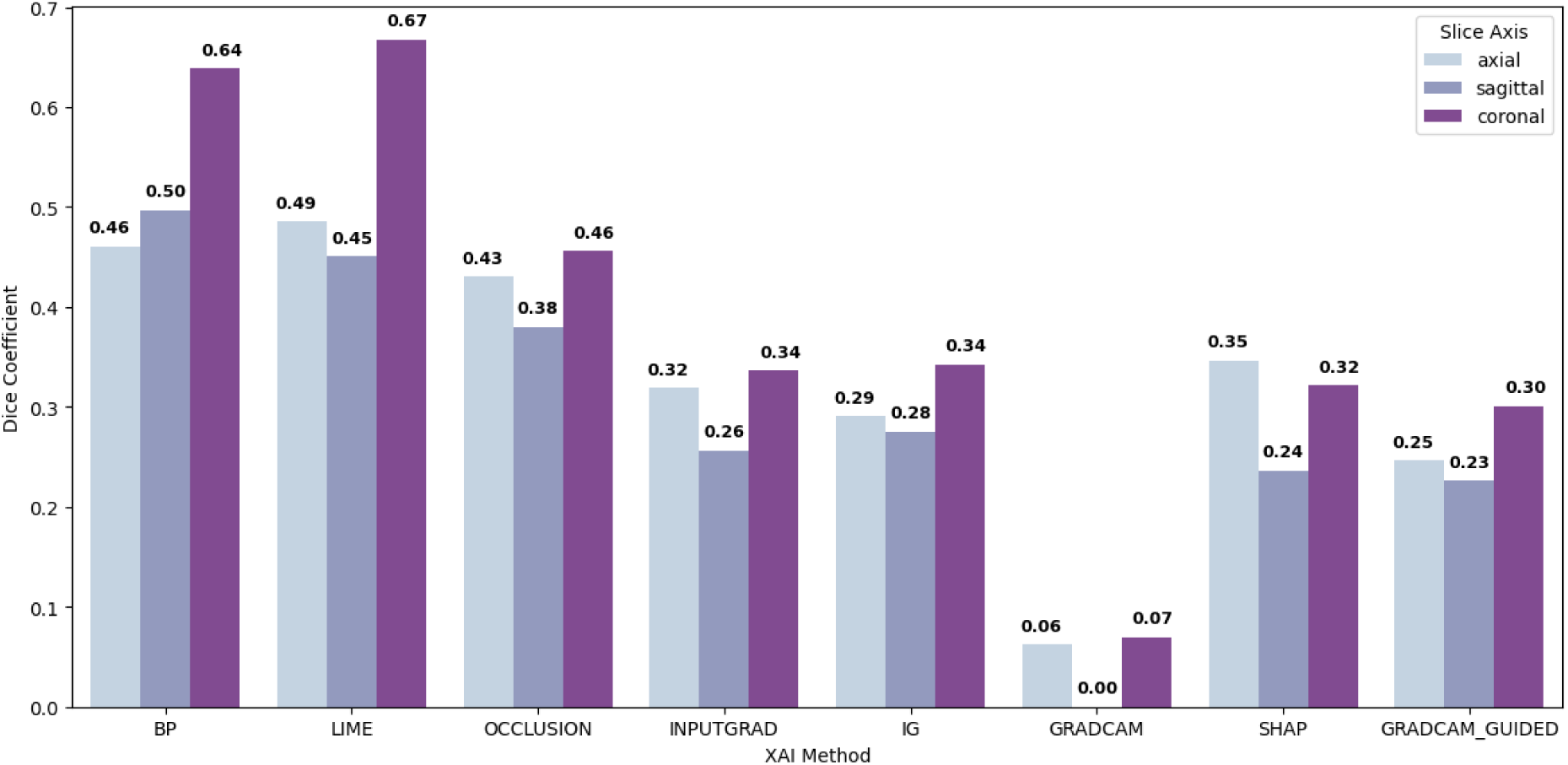
Dice coefficient between binarized saliency maps using the top 10% salient pixels/region in the average ‘AD’ heatmap across 8 XAI methods (only methods applicable to both architectures were included). Dice coefficients are given for models trained on three different MRI views: axial = light blue, sagittal = violet, coronal = purple. Abbreviations: ViT = vision transformer, -A = axial, -S = sagittal, -C = coronal, LRP = layer wise relevance propagation, AR = attention rollout, BP = backpropagation, IG= integrated gradients, INPUTGRAD = InputxGradient, CAM = class activation mapping, LIME = local interpretable model explanations, SHAP = (Shap)ley values.

**Figure 6.**
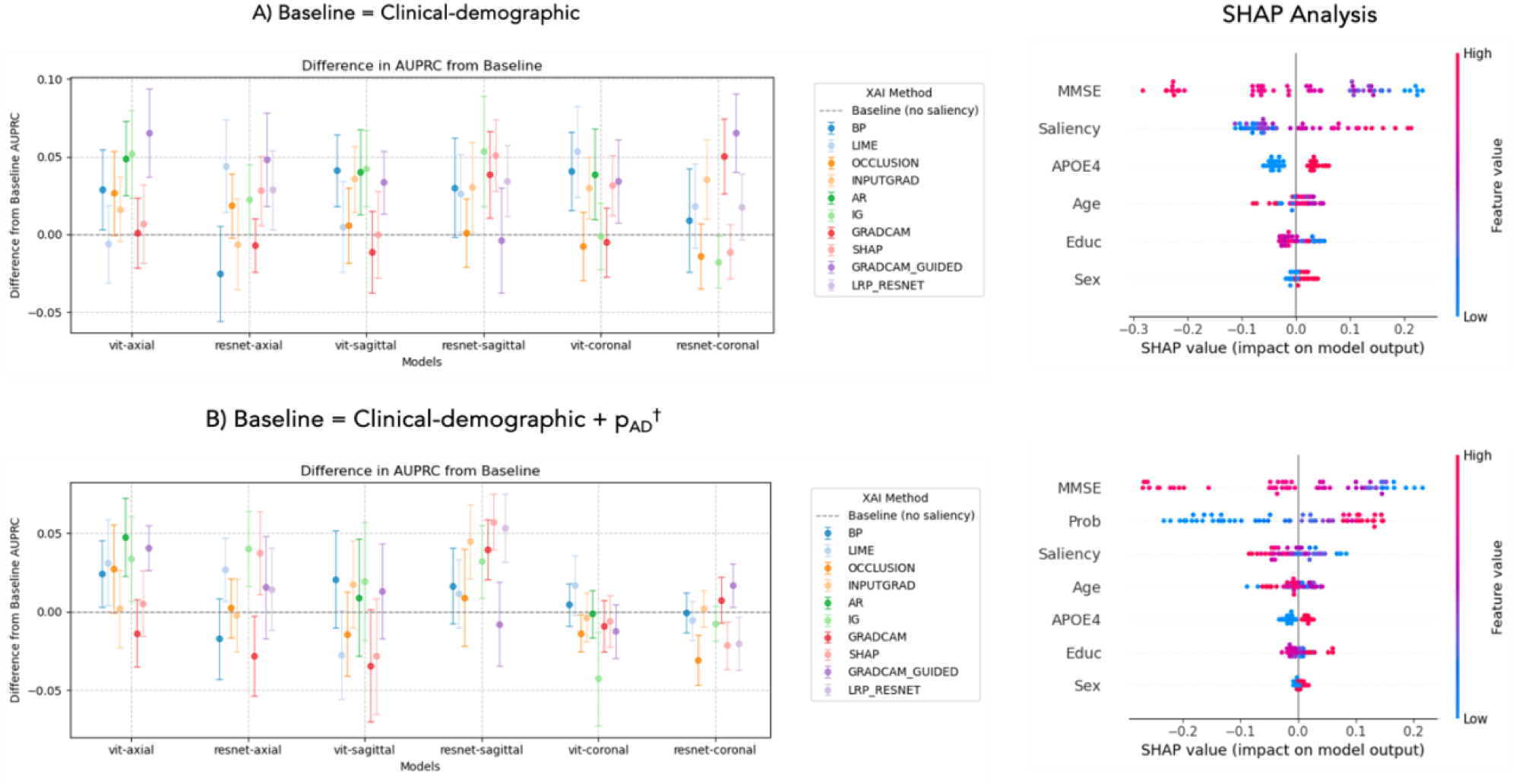
Difference in the area under precision-recall curve (AUPRC) across models where mean saliency is included in a Random Forest classifier trained and tested on MCI prognosis. We repeat this using 25 random seeds and plot the mean (points) and standard deviation (error bars). ★ denotes the model that yielded the best AUPRC value. SHAP summary plots indicate the contribution of each feature, shown for the optimal model. A) The difference is taken with respect to a model trained on clinical and demographic features only. B) The (relative) difference is taken with respect to a model trained on clinical-demographic features and corresponding image-based probabilities, p_AD_. Abbreviations: ViT = vision transformer, -A = axial, -S = sagittal, -C = coronal, LRP = layer wise relevance propagation, AR = attention rollout, BP = backpropagation, IG = integrated gradients, InputGrad = InputxGradient, CAM = class activation mapping, LIME = local interpretable model explanations, SHAP = (Shap)ley values.

### 4.3 Saliency maps for prognosis prediction

The results of using data from saliency maps to MCI to AD prognosis prediction are summarised in Figure 6 and Table 5. In Figure 6, we report the increase (mean and standard deviation) in AUPRC compared to a baseline where saliency is not included. For the clinical-demographic only analysis, the baseline AUPRC was 62.8% and is the same across all models. For baseline with both clinical-demographic features and imaging-based probability included, the difference is taken with respect to each model-specific baseline. SHAP analysis highlights the relative contribution of each feature and is computed for the optimal feature combination. In Table 5 we provide the performance metrics for Random Forest models trained using the optimal feature combinations and the two baseline models.

**Table 5.** Random Forest held-out test performance for MCI prognosis. Models were trained using clinical and demographic features, with the addition of mean saliency and the imaging-based probability, p_AD_. We report the mean and standard deviation of performance metrics obtained after 25 repeats. Abbreviations: demo = demographics, BACC = balanced accuracy, AUROC = area-under-the-receiver-operator-curve, AUPRC = area-under-the-precision-recall-curve, p_AD_ = imaging-based predicted probability of dementia. * Mean guided GradCAM saliency based on the axial ViT model (corresponds to ⭐ in Figure 8 6a). † pAD based on the coronal ViT model (baseline value with respect to ⭐ in Figure 6b). ♦ Mean LIME saliency and pAD based on the coronal ViT model (corresponds to ⭐ in Figure 6b)

|  | Prognosis (sMCI (25) vs. pMCI (31) – 25% all MCI participants) |  |  |
| --- | --- | --- | --- |
|  | BACC | AUROC | AUPRC |
| Clinical-demo | 59.2 ± 0.03 | 60.3 ± 0.01 | 62.8 ± 0.03 |
| Clinical-demo + saliency* | 62.9 ± 0.02 | 64.4 ± 0.02 | 69.4 ± 0.03 |
| Clinical-demo + p<sub>AD</sub><sup>†</sup> | 75.5 ± 0.03 | 78.5 ± 0.02 | 84.4 ± 0.02 |
| Clinical-demo + p<sub>AD</sub> + saliency<sup>♦</sup> | **76.6 ± 0.02** | **79.5 ± 0.02** | **86.1 ± 0.02** |

## 5 Discussion

This study explores the application of CNNs, ViTs and explainable XAI for advancing dementia diagnosis and prognosis. We show that pretrained ViTs can be effectively finetuned for AD classification and predicting MCI conversion, achieving strong performance even with 2D slices. We also propose novel approaches for multi-slice data fusion. Beyond performance, our work emphasizes the use of XAI to validate model behaviour and inform downstream tasks such as classification analysis. Our findings provide a comprehensive evaluation of current XAI approaches in the context of dementia prediction and valuable insights for future research.

### 5.1 Pretrained models can be effectively leveraged for dementia diagnosis and prognosis

For dementia classification, we obtained a balanced accuracy of 78.7% using mid-stack coronal slices compared to 83.4% with a fully 3D model. This difference was even smaller (3.6%) for MCI prognosis. These findings are pertinent to considerations around AI deployment in resource-constrained settings where access to high-resolution 3D data may be limited(41). We also assessed generalisability using held-out samples from NACC and an external ADNI cohort, addressing criticisms of existing studies that do not evaluate model transfer to unseen cohorts (3,4,42). Overall, 2D ResNet models slightly outperformed ViTs, which is likely due to the ViTs’ lack of intrinsic inductive biases and their reliance on patch-based attention for feature extraction(13). However, considering their fundamental modelling differences, our results raise questions about the trade-off between accuracy and interpretability, as ViTs’ lack of convolutional filters may make them more transparent.

To mitigate the limitations of single slices, we introduced three novel strategies for fusing information from orthogonal views. Surprisingly, jointly finetuning three ResNet networks allowed us to match the performance of a fully 3D network trained from scratch (+0.1% AUPRC, +2.2% AUROC, -2.75% BACC). Our approach aligns with other attempts to leverage ViTs in small datasets such as Alp and colleagues(43) who combined an ensemble and a recurrent network to capture inter-slice dependencies across 50 slices in each plane, achieving accuracies up to 99% on ADNI test data. However, our primary aim was not to optimise performance but to train models capable of generating meaningful explanations. Further work is needed to directly investigate the explainability of fusion models.

### 5.2 Explainable artificial intelligence as a sanity check

Model explanations can serve as a sanity check, verifying whether models focus on brain regions or show evidence of bias. Notably, to minimize preprocessing we chose not to apply skull stripping, resulting in non-zero non-brain MRI intensity which could introduce a source of bias. However, the saliency maps in Figure 3 confirm that the most salient regions were within the brain. Exceptions were the average GradCAM heatmap, where saliency outside the brain likely reflected artifacts from upsampling or the choice of convolutional layer, and also LIME and occlusion heatmaps, which are prone to noise due to their iterative mechanisms(19). Nonetheless, across most XAI methods we found that models converged upon brain regions known to be relevant to AD such as the hippocampus, cingulate gyrus and ventricular areas.

### 5.3 LIME and backpropagation appear robust to model architecture

A widely reported limitation of XAI is its sensitivity to biases in the underlying model. Some studies have quantified this sensitivity by randomizing layer weights and observing the effect on XAI outputs(5,6). In Figure 5 we assess the robustness of XAI outputs across CNN and ViT architectures by comparing the top salient pixels. These results indicate that backpropagation and LIME identified the most similar patterns across the two architectures with dice coefficients of 0.64 and 0.67 respectively in the coronal models. However it is important to consider that robustness in methods such as GradCAM that are directly related to model outputs will be also be affected by differences in predicted probabilities (Supplementary Figure A3).

### 5.4. Saliency maps enhanced MCI prognosis

GradCAM mean saliency from the axial ViT model improved the AUPRC by 6.6% compared to a clinical-demographic baseline. Including both LIME mean saliency and predicted probabilities from the coronal ResNet improved AUPRC by a further 16.7%. SHAP analysis revealed that MMSE was the driving feature but saliency ranked highly in both cases.

However, high guided GradCAM mean saliency had a positive effect, whilst high mean LIME saliencies had negative SHAP values. This divergence may be linked to differences in XAI mechanisms, as perturbation-based methods like LIME are more reflective of change to the model output rather than the value itself. This highlights the importance of understanding the assumptions behind XAI methods to interpret their outputs accurately(3). We also note that in some cases including mean saliency hindered performance, potentially due to polarizing feature influences or poor-quality heatmaps. Despite this, our results illustrate how XAI outputs can provide valuable insights for downstream tasks like classification analysis, offering additional information beyond traditional clinical markers.

### 5.5 Study limitations and future directions

A key limitation of our study is that we have restricted our analysis to 2D models. The high computational cost of training 3D models and certain XAI techniques, emphasises the significance of achieving competitive performance with single (or few) slices. Our results also contrast with a recent study which reported superior performance of ViTs over ResNets in dementia classification(26). However conducting a fair head-to-head comparison of these architectures is not trivial despite our efforts to keep design choices such as training data size and strategies consistent(44). More importantly, we re-empathise that performance was not the focus and opportunities to optimise model hyperparameters further may affect our results. Future studies could explore extensions to the ViT that are still convolution-free, such as the Swin Transformer, which may balance performance and interpretability. We also found that models trained on coronal slices performed best, which is likely due to the prominence of the hippocampi in this plane. As such our approach could be extended by targeting specific slice locations to increase the discriminative ability without requiring the entire 3D volume.

Also, our comparison of the different XAI outputs remains largely qualitative due to the absence of a ground-truth. Existing XAI validation studies have used ROIs derived from meta-analyses(45,46), or simulated perturbations to capture sensitivity to specific features^47^(47). Whilst these approaches can be useful for group averages, there is still a need to design strategies to validate saliency at the individual level.

There are also limitations related to the data used. Although we applied a weighted loss function to discourage the model from focusing on the majority class, we highlight that class imbalance will affect the interpretation of performance metrics. Therefore we report the balanced accuracy, AUROC and AUPRC, to provide a fuller picture. The limited availability of pretrained ViT models restricted us to those pretrained on natural images. In the advent of foundation models and open, medical imaging datasets, further work could investigate the use of brain-specific pretraining tasks such as brain-age prediction(25) or sex classification(23).

Uncertainty introduces another challenge, as potential misdiagnoses and delays between imaging and clinical assessment may impact the reliability of assigned labels. To mitigate this, we used clinical assessments made within 6 months of the baseline scan, however as the ADNI visits are more routinely collected, this difference could explain the increase in performance in external test data. For MCI participants we used a window of 3-4 years for conversion to align closely with ADNI. However, ADNI MCI participants had a higher MMSE compared to NACC on average, indicating a healthier cohort that may be harder to classify. We also observed lower zero-shot prognostic recall in ADNI (highest value of 45%) compared to NACC (highest value of 73%), which could be linked to scan quality differences (Supplementary Figure A2).

Many XAI methods rely on parameter choices that can affect their results. For example, the occlusion value is a key parameter, and while we used a default value of 0, this value can carry information in the context of MRI. Similarly, our modified LRP implementation for ResNets relied on predefined rules for specific layers, which were chosen to align with the original implementation(48). Exploring alternative configurations may yield different insights and warrants further investigation.

### 5.6 Implications for clinical translation

This study serves as a starting point for understanding the applicability of XAI to dementia prediction tools. We suggest that due the high variability of XAI outputs, future work should consider multiple XAI methods to derive insights on model behaviour and detect potential biases. We also emphasize that the performance advantages of CNNs should be weighed against the potential transparency of ViTs, particularly in clinical applications. The granularity of salient regions varied markedly across XAI methods, due to their underlying assumptions and mechanisms. Whilst we found that models converged on the most salient regions after thresholding, these differences may be significant depending on the task and intended use of model explanations. For example, occlusion and LIME outputs were more brain-like in size and shape, which may be more useful for visually identifying known regions of interest. In contrast, gradient-based techniques such as Integrated Gradients highlight pixel-level saliency and may be better suited systematic evaluations of robustness, or applications where local accuracy is important such as segmentation or voxel-based analyses.

### 5.7 Conclusion

We present a comprehensive comparison of several explanation methods across both CNN and ViT architectures. We demonstrate that transfer learning can be used to leverage pretrained models, exploring strategies for fusing information across multiple views. Despite their limitations, XAI methods provide useful information, serving as a tool to detect bias and to understand what information the model is focusing on.

## Author contributions

SAM: Study design, implementation, materials and data collection, visualisation and initial manuscript preparation. AZ: Study design (model training). JQ: Material and data collection. PI and AI: Study design (analysis). FB: Study conception, study design and methodology. JHC: study conception, study design, methodology. All authors read, reviewed and approved the final manuscript.

## Data Availability

All data produced are available online at https://adni.loni.usc.edu and https://naccdata.org.

https://naccdata.org

## Acknowledgements

This work is supported by the Engineering and Physical Sciences Research Council funded Centre for Doctoral Training in Intelligent, Integrated Imaging in Healthcare (i4health) (EP/S021930/1) and the Department of Health’s NIHR-funded Biomedical Research Centre at University College London Hospitals. Phoebe Imms is supported by a USC/UCLA Biodemography Center Pilot Project Award through a grant from the National Institute on Aging (P30AG017265), and a T32 Multidisciplinary Training in Gerontology fellowship (2T32AG000037-46). Andrei Irimia acknowledges support from the National Institutes of Health (NIH) under grants R01 NS 100973, RF1 AG 082201, and R01 AG 079957, from the Department of Defense under contract W81XWH-18-1-0413, from anonymous donors, from the Hanson-Thorell Research Scholarship Fund, the Undergraduate Research Associate Program (URAP), and the Center for Undergraduate Research in Viterbi Engineering (CURVE) at the University of Southern California.

The NACC database is funded by NIA/NIH Grant U24 AG072122. NACC data are contributed by the NIA-funded ADRCs: P30 AG062429 (PI James Brewer, MD, PhD), P30 AG066468 (PI Oscar Lopez, MD), P30 AG062421 (PI Bradley Hyman, MD, PhD), P30 AG066509 (PI Thomas Grabowski, MD), P30 AG066514 (PI Mary Sano, PhD), P30 AG066530 (PI Helena Chui, MD), P30 AG066507 (PI Marilyn Albert, PhD), P30 AG066444 (PI David Holtzman, MD), P30 AG066518 (PI Lisa Silbert, MD, MCR), P30 AG066512 (PI Thomas Wisniewski, MD), P30 AG066462 (PI Scott Small, MD), P30 AG072979 (PI David Wolk, MD), P30 AG072972 (PI Charles DeCarli, MD), P30 AG072976 (PI Andrew Saykin, PsyD), P30 AG072975 (PI Julie A. Schneider, MD, MS), P30 AG072978 (PI Ann McKee, MD), P30 AG072977 (PI Robert Vassar, PhD), P30 AG066519 (PI Frank LaFerla, PhD), P30 AG062677 (PI Ronald Petersen, MD, PhD), P30 AG079280 (PI Jessica Langbaum, PhD), P30 AG062422 (PI Gil Rabinovici, MD), P30 AG066511 (PI Allan Levey, MD, PhD), P30 AG072946 (PI Linda Van Eldik, PhD), P30 AG062715 (PI Sanjay Asthana, MD, FRCP), P30 AG072973 (PI Russell Swerdlow, MD), P30 AG066506 (PI Glenn Smith, PhD, ABPP), P30 AG066508 (PI Stephen Strittmatter, MD, PhD), P30 AG066515 (PI Victor Henderson, MD, MS), P30 AG072947 (PI Suzanne Craft, PhD), P30 AG072931 (PI Henry Paulson, MD, PhD), P30 AG066546 (PI Sudha Seshadri, MD), P30 AG086401 (PI Erik Roberson, MD, PhD), P30 AG086404 (PI Gary Rosenberg, MD), P20 AG068082 (PI Angela Jefferson, PhD), P30 AG072958 (PI Heather Whitson, MD), P30 AG072959 (PI James Leverenz, MD).

Data collection and sharing for the Alzheimer’s Disease Neuroimaging Initiative (ADNI) is funded by the National Institute on Aging (National Institutes of Health Grant U19AG024904). The grantee organization is the Northern California Institute for Research and Education. In the past, ADNI has also received funding from the National Institute of Biomedical Imaging and Bioengineering, the Canadian Institutes of Health Research, and private sector contributions through the Foundation for the National Institutes of Health (FNIH) including generous contributions from the following: AbbVie, Alzheimer’s Association; Alzheimer’s Drug Discovery Foundation; Araclon Biotech; BioClinica, Inc.; Biogen; Bristol-Myers Squibb Company; CereSpir, Inc.; Cogstate; Eisai Inc.; Elan Pharmaceuticals, Inc.; Eli Lilly and Company; EuroImmun; F. Hoffmann-La Roche Ltd and its affiliated company Genentech, Inc.; Fujirebio; GE Healthcare; IXICO Ltd.; Janssen Alzheimer Immunotherapy Research & Development, LLC.; Johnson & Johnson Pharmaceutical Research & Development LLC.; Lumosity; Lundbeck; Merck & Co., Inc.; Meso Scale Diagnostics, LLC.; NeuroRx Research; Neurotrack Technologies; Novartis Pharmaceuticals Corporation; Pfizer Inc.; Piramal Imaging; Servier; Takeda Pharmaceutical Company; and Transition Therapeutics.

## Consent statement

Consent was not necessary for this work.

