## Supplementary Material for "Explainable artificial intelligence for neuroimaging-based dementia diagnosis and prognosis"

**A1 –** Participant filtering and data processing pipeline for NACC

Images were downloaded from the NACC June 2022 freeze (prior to SCAN initiative availability). Therefore, all imaging data is voluntarily uploaded by the individual ADRCs and are not subject to routine preprocessing or quality control checks as in the ADNI database. To remove poor quality scans, we utilised visual assessment and MRIqc image quality metrics to exclude images with any outliers based on CJV, SNR or CNR values. Additionally, participants were removed due to download issues, incorrect `nifti` header information and a lack of available clinical diagnosis of CN, MCI or ADD within a 6-month time-period from their baseline scan. Some participants who received follow-up diagnosis were classified as reverters where their baseline diagnosis of MCI or AD later improved (e.g., MCI to CN, AD to MCI, or AD to CN). These participants were also removed from our analysis.

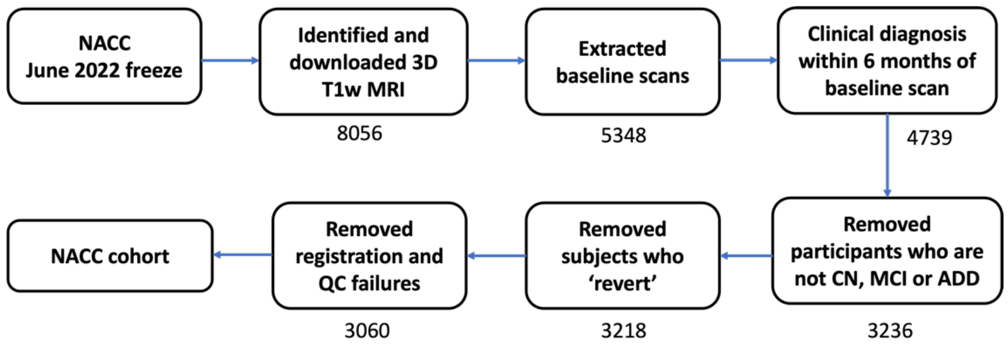


Figure A1. Data flow for NACC data processing.

**A2 –** MRIqc image quality metrics in NACC and ADNI

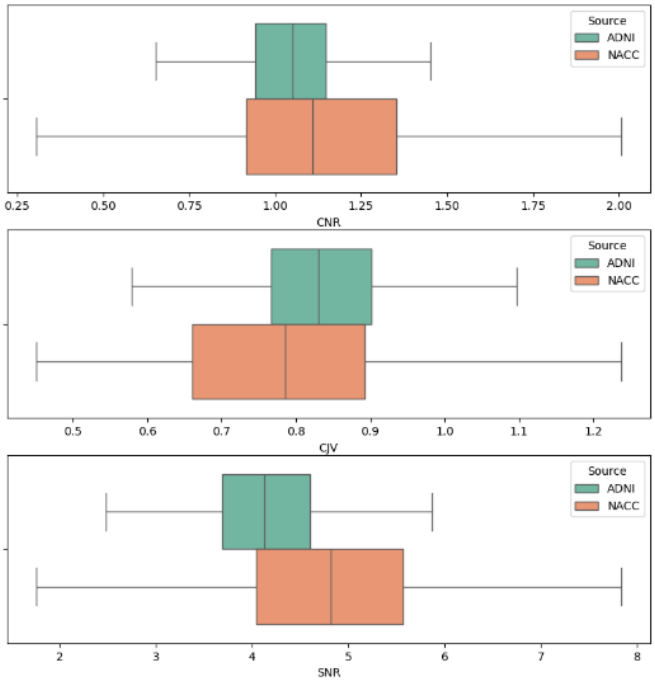


Figure A2. MRIqc derived image-quality metrics for NACC and ADNI 3D T1-weighted MRI data (AD and CN individuals only, after removal of outliers). Note, the two datasets have different sample sizes.

**A3 –** CNN and ViT predicted probabilities

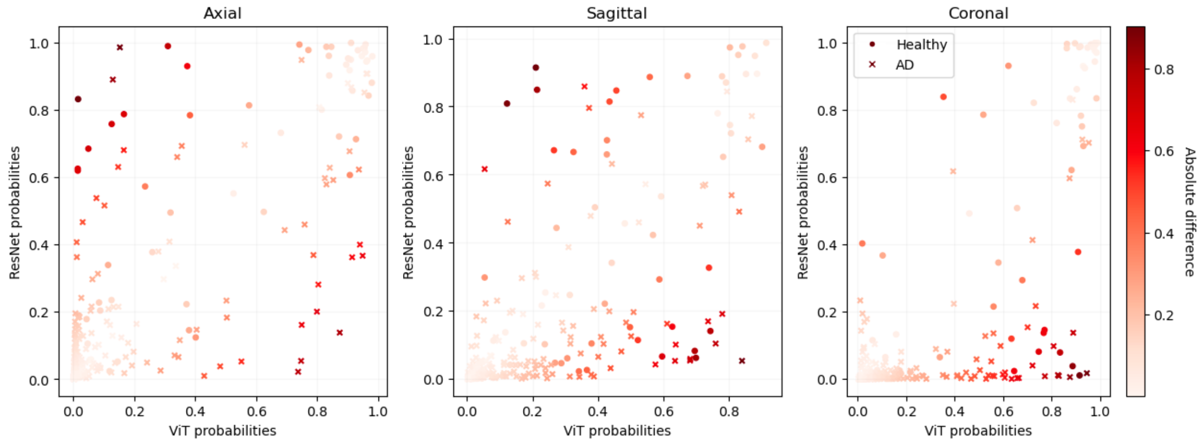


Figure A3. Scatter plots of single-slice model test predictions for AD classification across both architectures. Models tended to agree on difficult to classify cases.

**A4 –** Non-brain saliency across XAI methods and models

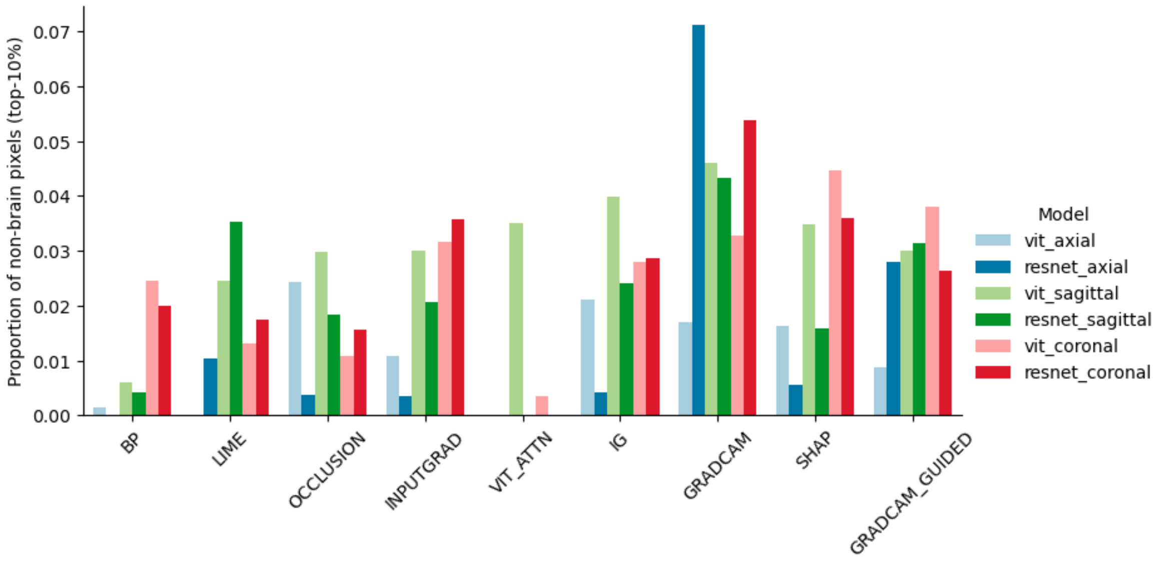


Figure A4. Proportion (%) of pixels (in top 10% percentile) that lie outside of brain across XAI methods and slice-wise models. Abbreviations: ViT = vision transformer, LRP = layer wise relevance propagation, AR = attention rollout, BP = backpropagation, IG = integrated gradients, INPUTGRAD = InputxGradient, CAM = class activation mapping, LIME = local interpretable model explanations, SHAP = (Shap)ley values.

**A5 –** Mean ‘CN’ heatmap across XAI methods and models

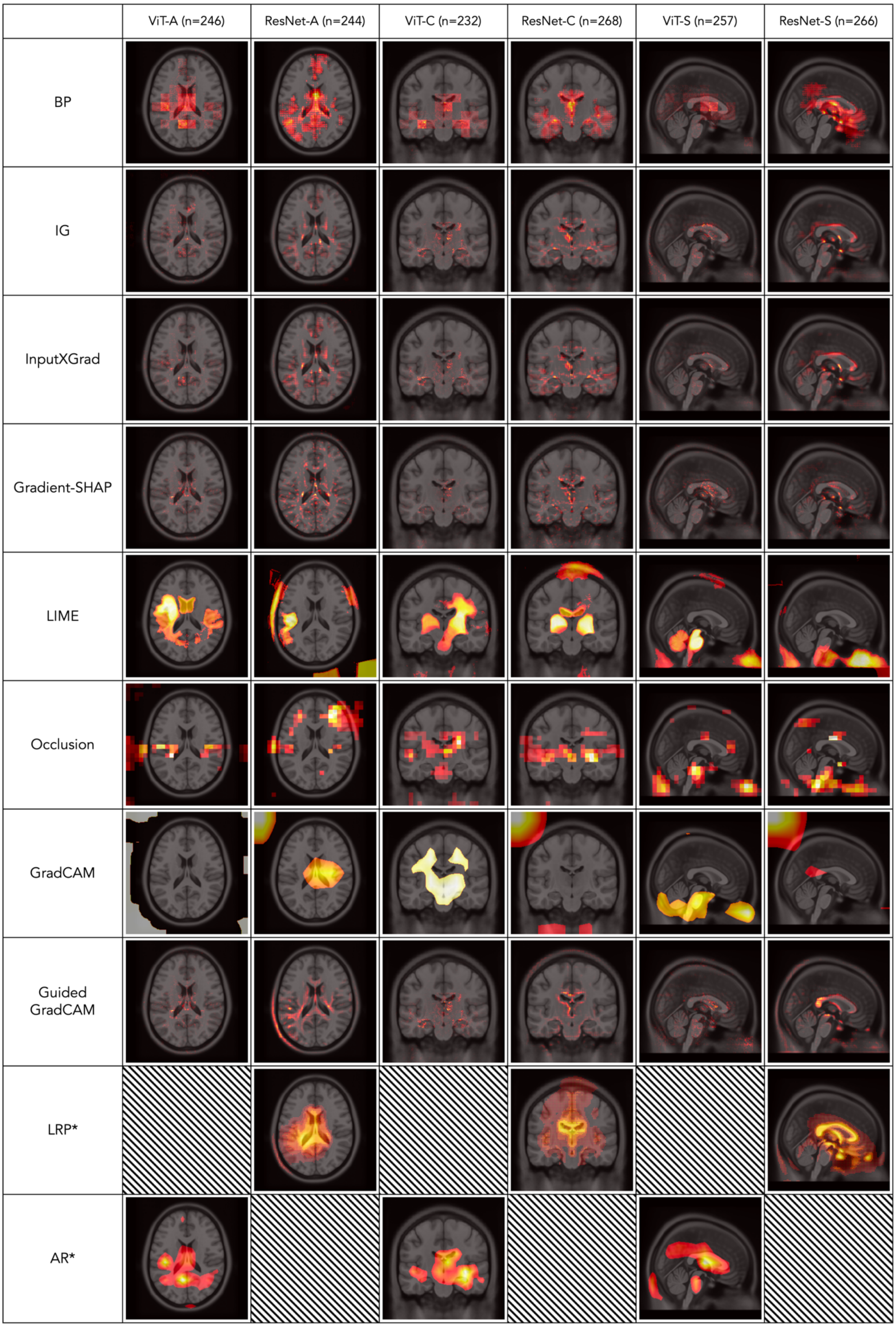

Figure A5. Top 10% of pixel intensities for mean heatmaps across ten XAI methods (averaged over true negative and false negative model predictions with the corresponding number of participants given in brackets. Abbreviations: ViT = vision transformer, -A = axial, -S = sagittal, -C = coronal, LRP = layer wise relevance propagation, AR = attention rollout, BP = backpropagation, IG = integrated gradients, INPUTGRAD = InputxGradient, CAM = class activation mapping, LIME = local interpretable model explanations, SHAP = (Shap)ley values. * Denotes methods that were not implementable for both architectures.

**A6 –** Random Forest hyperparameters

| Parameter | Search values |
|---|---|
| **max_depth** | [None, 2, 5,10] |
| **min_samples_leaf** | [1,3,5,10] |
| **min_samples_split** | [2,5,10] |
| **n_estimators** | [1,3,10,50] |
| **criterion** | [‘gini’, ‘entropy’, ‘log_loss’] |
| **class_weight** | [‘balanced’, None] |

Table A6. Random Forest hyperparameters and values that were assigned during 5-fold CV grid search optimisation using scikit-learn.

**A7 –** Vision transformer and ResNet architecture details and hyperparameters

| | **Vision Transformer** | **ResNet** |
|---|---|---|
| Architecture | *vit-tiny-patch16-224* | *resnet18d_21k* |
| # Params | 5.7M | 11.7M |
| Fixed parameters (which differ from default) | in_chans: 1<br>attn_dropout: 0.1<br>num_classes: 1 | in_chans: 1<br>num_classes: 1 |
| Hyperparameter search space | Initial learning rate: [1e-10, **1e-4**, 1e-2]<br>Scheduler: **[‘**reduce_on_plateau’, **None**]<br>Optimiser: [‘sgd’, **‘adamw’**]<br>Dropout: [0, 0.1, **0.5**]<br>Momentum: [**0.9**, 0.5, 0.99]<br>Weight decay: [0, 1e-2, **1e-4**] | Initial learning rate: [1e-10, 1e-4, **1e-2**]<br>Scheduler: [**‘reduce_on_plateau’**, None]<br>Optimiser: [**‘sgd’**, ‘adamw’]<br>Dropout: [**0**, 0.1, 0.5]<br>Momentum: [0.9, 0.5, **0.99**]<br>Weight decay: [0, 1e-2, **1e-4**] |

Table A7. Details on the model architectures used for the ViT and ResNet-18 and hyperparameter search values (optimised values are shown in **bold**). We performed hyperparameter search for axial slices (and 3D) models only, and kept these parameters fixed for the remaining models. Pretrained models were obtained from the `timm` library.

**A8 –** MLP hyperparameters for late fusion model

| Parameter | Search values | Method |
|---|---|---|
| **hidden_size** | [1, 3, 5] | Grid search |
| **num_layers** | [0,1,3] | Grid search |
| **learning_rate** | [0.0001, 0.001, 0.01, 0.1] | Random choice |
| **batch_size** | [8,32,64] | Random choice |

Table A8. Multi-layer perceptron hyperparameters and values that were explored to optimise triplanar ensemble (late) models over ten trials. The optimal model was chosen based on the validation performance (average precision) using NACC-trained ViT model predictions and kept fixed for all other triplanar-late models.
